# Instability of Global Burden of Disease Estimates of Deaths and DALYs from Major Risk Factors

**DOI:** 10.1101/2025.11.14.25340145

**Authors:** Emmanuel A. Zavalis, Angelo Maria Pezzullo, John P.A. Ioannidis

**Author notes:** Correspondence to: John P.A. Ioannidis, MD, DSc, 3180 Porter Drive, Room A129, Stanford Research Park, Palo Alto, CA 93404, USA. **Disclosures:** This study is independent of the Global Burden of Disease (GBD) enterprise and the Institute for Health Metrics and Evaluation (IHME). The authors used only publicly available, published GBD results and did not access any non-public data, software, or pre-publication materials. IHME/GBD had no role in the conception, design, data collection, analysis, interpretation, writing, or decision to submit. AMP is a member of the GBD Collaborator Network (non-financial relationship); no financial conflicts to declare. The views expressed are solely those of the authors and do not represent IHME, the GBD enterprise, the University of Washington, or any funders. **Funding:** none. **Data availability:** All data are publicly available.

## Abstract

**Importance:** The Global Burden of Disease (GBD) study provides widely used estimates of mortality and disability-adjusted life years (DALYs) attributable to risk factors.

**Objective:** To evaluate the variability and consistency of GBD risk factor estimates for mortality and DALYs.

**Data Sources:** GBD Risk Factor collaboration estimates extracted from published tables and IHME repository.

**Study Selection:** GBD Risk Factor collaboration publications (2010-2023).

**Data Extraction and Synthesis:** Death and DALY estimates were manually extracted by one reviewer with independent validation of a random sample of 100 by another with no discrepancies. Risk factor naming was harmonized across iterations to ensure comparability; those with inconsistent definitions were excluded.

**Main Outcomes and Measures:** We calculated the fluctuations in deaths and DALYs for each risk factor across GBD iterations for different years (2010-2023), and between the original and subsequently revised estimates for each year (1990-2021) expressed in the ratio of the min-max range to the mean (R/M) and coefficient of variation (CV). We examined in more detail analyses diet and low physical activity. Finally, point estimates were compared to the previous iterations’ estimates 95% uncertainty intervals (95%UI) for GBD 2019, 2021 and 2023.

**Results:** Across GBD iterations from 2010 to 2023, the median R/M was 0.8 (range, 0–3.8) for deaths, and 0.7 (range, 0.1–3.3) for DALYs. Among level 2 dietary and child and maternal malnutrition death estimates showed high variability (R/M>1 for 7/16 and 3/8 of risks, respectively). When comparing original estimates with GBD 2019, 2021, and 2023 estimates for the same years, the median R/M was 0.5 (0.0-2.9) for deaths and 0.4 (0-2.9) for DALYs. The CV was above 0.2 for 320/580 (52%) of death and 306/609 of DALY estimates. 70-96% of point estimates for red meat, sugar-sweetened beverages, fruits, vegetables and seafood omega-3 fatty acids in GBD 2021 fell outside the GBD 2019 95%UI. In GBD 2023, only diet high in trans fats had over half of point estimates outside the GBD 2021 95%UI.

**Conclusions and Relevance:** GBD estimates show large instability, particularly for behavioral risks, making them unlikely to simply reflect genuine changes over time, and warranting caution in interpretation.

**Key Points:** *Question:* How stable and consistent are the Global Burden of Disease (GBD) estimates for mortality and disability-adjusted life years (DALYs) attributable to major risk factors across iterations from 2010 to 2023?

*Findings:* In this study comparing estimates across eight GBD iterations, substantial variability was observed. Behavioral, particularly dietary risks showed the greatest instability. Comparing revised estimates across iterations half the estimates had a coefficient of variation exceeding 0.2. A third of estimates for dietary risks in GBD 2021 fell outside the corresponding GBD 2019 uncertainty intervals.

*Meaning:* GBD risk factor estimates, especially for behavioral and dietary risks, show marked inconsistency likely reflecting methodological or data changes rather than true burden shifts.

## Introduction

The Global Burden of Disease (GBD) study was initiated more than three decades ago^1^, and has provided regularly updated iterations over time with the most recent covering estimates for 2023^2,3^. GBD estimates of the relative impact of diseases and risk factors on deaths and disability-adjusted life years (DALYs) are a widely used and cited resource in global health and epidemiology. GBD collaborators synthesize evidence and use sophisticated modeling of risk and prevalence data to generate these estimates that then inform policy, healthcare professionals and the general public^4,5^. Each GBD iteration not only estimates the current burden, but also retrospectively models and revises estimates for prior years. E.g., GBD 2010 included estimates of deaths due to smoking for both 1990 and 2010^6^.

Given its scientific and policy influence, the reliability of these estimates are crucial. However, risk factor epidemiology is inherently complex, and results may vary widely depending on both genuine differences and biases in different studies and calculations. Establishing both causality and the magnitude of effect is difficult, even for exposures with known health effects^7^. Capturing long-term outcomes requires follow-up hard to justify in randomized trials, while observational studies suffer many biases. Randomized trials are unethical for harmful factors (unless they address interventions to potentially reduce them). Mendelian randomization studies are sensitive to pleiotropy and population stratification, and strong genetic instruments are often lacking^8^.

Overall, the reliability of risk factor epidemiology is often limited—particularly for exposures like diet, which are difficult to quantify and intervene upon^9^. Modeling these factors globally adds another layer of difficulty, given the fragmentary and potentially biased nature of data worldwide. Previous critiques of the GBD Risk Factor estimates have focused on methodological decisions, e.g. thresholds used to define risk groups for low physical activity and dietary risks^9–11^. Given the scope of the GBD initiative, understanding the robustness of each assumption and of each piece of data involved in the modeling is challenging. However, as GBD collaborators update their estimates regularly, the overall stability and consistency of reported estimates can be assessed. The reported (updated and revised) estimates reflect the end product of all changes that happen in the GBD input data, including assumptions, and calculations, and they are the numbers that eventually become influential for policy decisions.

A fundamental question is whether these estimates remain relatively steady or change markedly over time. Some changes over time may occur because of genuine decreases (e.g. progress in reducing the prevalence of risk factors or finding better interventions to diminish their effects) or genuine increases (e.g. wider spread of harmful factors across populations). However, for most risk factors such changes may not be so prominent over the relatively short period from 2010 to 2023, where eight different iterations of GBD estimates are available. Unstable estimates for the same risk factor over these iterations may instead mostly reflect the impact of imperfect data sources and of modeling choices. Therefore, through quantifying the instability of GBD estimates empirically we aim to indirectly probe the overall reliability of the estimates. We also aim to understand whether some risk factors have witnessed much larger fluctuations in their estimates compared to others.

## METHODS

The study is a meta-epidemiological assessment. There is no reporting guideline directly pertinent to this specific design, but we used whatever items were pertinent from the Preferred Reporting Items for Systematic reviews and Meta-Analyses^13^. Our analyses are exploratory and no protocol was pre-registered. Our analyses including GBD iterations until 2021 were completed by the fall of 2025; then they were updated to include also 2023 estimates as these became available in October 2025.

### GBD Data

Mortality and disability-adjusted life-year (DALY) estimates were extracted by EAZ from the 2010, 2013, 2015, 2016, 2017, 2019, 2021, and 2023 iterations of the Global Burden of Disease (GBD) study^2,6,14–17^ (Table 1). Estimates from GBD 2010 through 2017 were manually retrieved from published summary tables, whereas GBD 2019, 2021 and 2023 data were downloaded from the Institute for Health Metrics and Evaluation (IHME) online repositories^18–20^. A random sample of 100 manually extracted entries was cross-validated by AMP with no discrepancies identified.

**Table 1.**
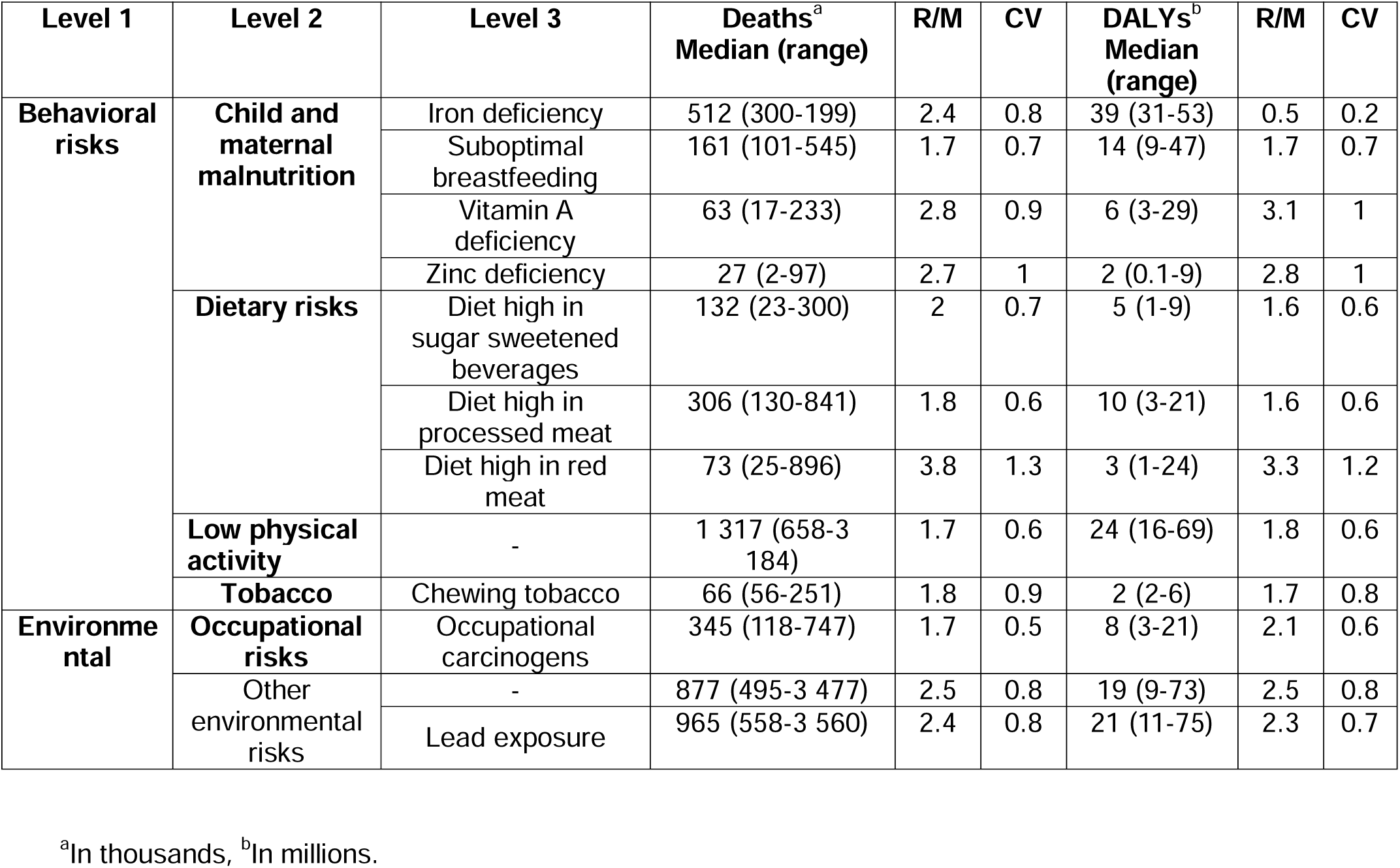
Level 2 risks with range to median ratio R/M > 1.5 for deaths or DALYs.

Two analytic datasets were constructed (eTable 1):

- Dataset 1: Included death and DALY estimates from each GBD iteration (2010–2017) and overlapping estimates as re-published in GBD 2019 and 2021 for years previously estimated (2010, 2013, 2015, 2016, 2017) as well as the original 2019 and 2021 estimates.
- Dataset 2: Included the full time series for dietary risks (1990–2021) estimated in GBD 2019, 2021 and GBD 2023, stratified by sex and geography (World Bank regions), as well as aggregated totals for major causes (non-communicable diseases, communicable diseases).

### Risk Factor Classification and Harmonization

Risk factors were categorized according to the GBD hierarchical framework (Levels 1–3), with Level 1 representing broad categories (behavioral, metabolic, environmental/occupational), Level 2 capturing subgroups (e.g., Tobacco, Dietary Risks), and Level 3 specifying individual risks (e.g., Diet high in sodium). Terminology was standardized across iterations to ensure comparability (eMethods). Risk factors with estimates available only in a single iteration were excluded. Level 4 risks were not analysed for ease of presentation and to focus on the most impactful aggregated risks.

### Outcomes and Statistical Analysis

We first described absolute fluctuations in deaths and DALYs across GBD iterations for the year of analysis (i.e., GBD 2010 estimates for 2010, GBD 2013 estimates for 2013, GBD 2015 estimates for 2015, etc.). These fluctuations were then summarized using the min-max range-to-mean ratio (R/M) and the proportion of estimates within each risk level with R/M exceeding 1.

Secondly, we analyzed the differences in the estimates for the same year, including the original estimate (e.g. GBD 2015 estimate for 2015) and any subsequent revisions published in later years (e.g. GBD 2019 estimate for 2015 and GBD 2021 estimate for 2015). Here, the only differentiating factor was the original versus revised iterations of analysis. We also calculated the coefficient of variation for these to provide an interpretation of the variability where the differentiating factor was the GBD iteration.

Third, dietary and low physical activity risks, which have been subject to debate^10–12^, were examined in greater detail through graphical presentation of absolute estimates and rankings.

Finally, for dietary and low physical activity risks, we compared the GBD 2021 point estimates with the 95% uncertainty intervals (UIs) reported in GBD 2019 (eFigure 1); and the GBD 2023 estimates with 95% UIs from GBD 2021.

All code and data are publicly available at https://github.com/zavalis/pubs_meta/tree/main/gbd%20variability.

## RESULTS

### Analyzed risk factors

After harmonization of risk factor naming, 101 unique risk factors were obtained. Of these, 95 were estimated across more than one iteration. Following exclusion (see eMethods for harmonization) of 23 Level 4 risk factors (primarily occupational risks), 5 risk factors related to childhood sexual abuse and bullying, and the combined ‘Alcohol and drug use’ Level 2 risk, 66 risks remained for the present analysis. Of these, one risk factor was Level 0, three were Level 1, 19 were Level 2, and 43 were Level 3 (eTable 2).

### Level 1 and 2 risk factors: estimates across GBD iterations, 2010 to 2023

Across successive GBD iterations, estimates of deaths and DALYs attributable to major risk factor categories showed a general increasing trend, particularly for metabolic and environmental/occupational risks, while estimates for behavioral risks display an oscillating behavior (Figure 1). Total deaths attributed to all risk factors rose from 30.8 million in GBD 2013 to 34.8 million in GBD 2023. Deaths attributable to metabolic risks increased steadily over the same period, from 15.7 million to 19.0 million, and environmental and occupational risks rose from 8.2 million to 13.9 million. In contrast, behavioral risk-related deaths increased initially but returned to earlier levels by 2021. DALY estimates broadly followed similar patterns, with metabolic and environmental risks showing sustained increases, while behavioral risks fluctuated.

**Figure 1.**
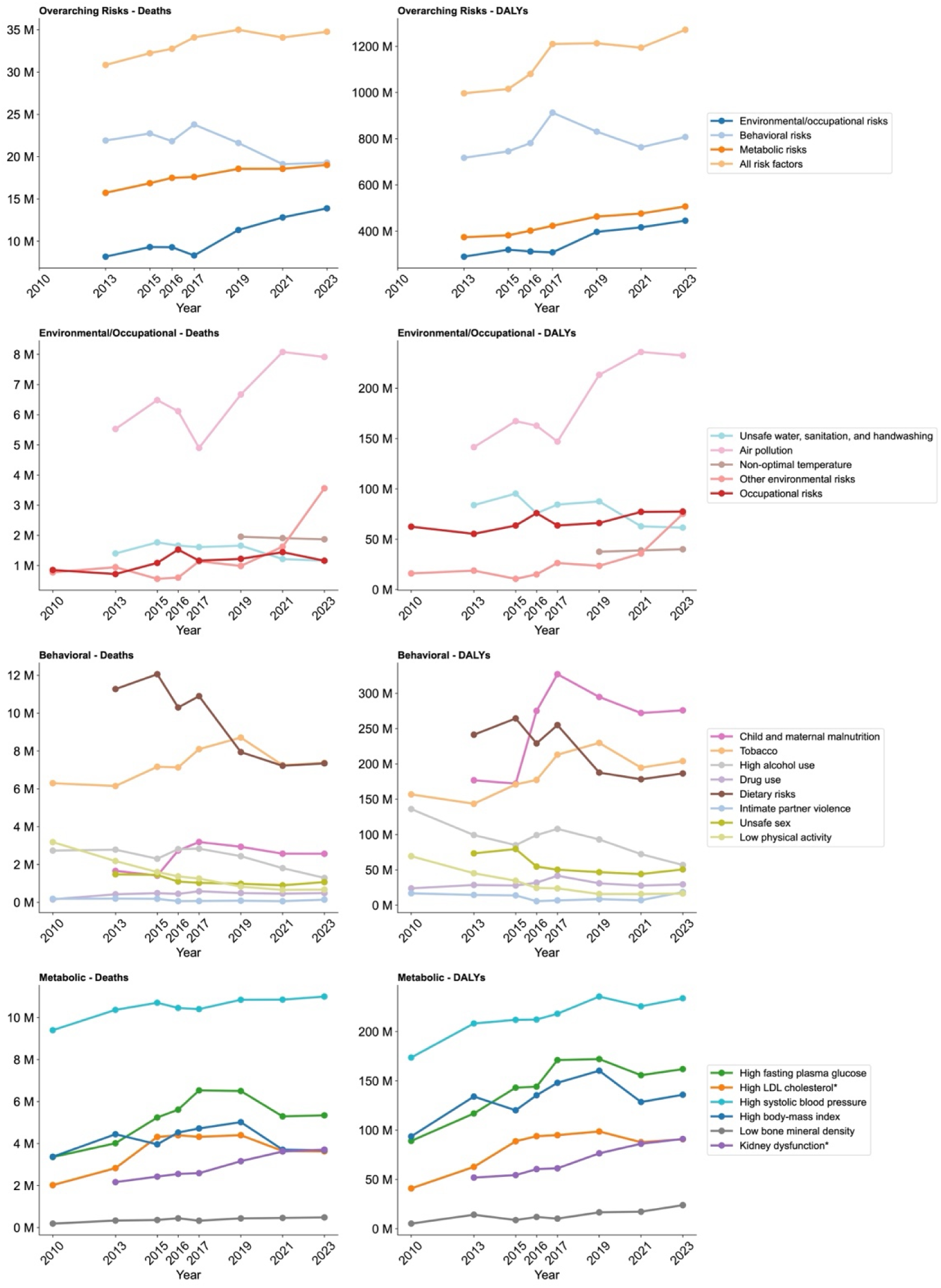
Estimated total deaths and DALYs attributable to Level 1 risk factors (Overarching risks), and to Environmental/Occupational, Behavioral and Metabolic Level 2 risks across GBD iterations.

At the Level 2 risk factor level, variability was more pronounced. Deaths attributed to air pollution increased from 4.9 million to 8.1 million across three GBD iterations (2017 to 2021) and in 2023 to 7.9 million, whilst dietary risks showed a decline in estimated deaths, from 11.3 million in GBD 2013 to 7.9 million by GBD 2019 stabilising in GBD 2021 and GBD 2023 to 7.3 million. Deaths linked to low physical activity declined gradually across all iterations from 2.2 million to 670 thousands. Fluctuations were also evident among the Level 2 risk DALY estimates. Air pollution saw a 66.3 million increase in attributed DALYs between GBD 2017 and 2019. Over the same interval, dietary risks declined by 67.3 million DALYs. Notably, DALY estimates for child and maternal malnutrition—unlike for most other behavioral risks—increased by over 102.9 million DALYs between GBD 2015 and 2016.

### Level 3 risk factors

Fluctuations amongst Level 3 risk factors were evident across environmental, behavioral, nutritional, and occupational exposures (eFigure 2). Particulate matter pollution estimates showed the largest increase, with deaths rising by 3.2 million (from 4.6 to 7.8 million) and DALYs by 88.5 million (from 143.0 to 231.5 million) from 2017 to 2021. However, occupational risk estimates remained relatively stable except for occupational injuries that successively increased from 311 thousand in 2019 to 550 thousand in 2023. Lead exposure saw a marked increase from 1.5 million to 3.5 million deaths between 2021 and 2023. Among tobacco-related risk factors, smoking death estimates increased from 5.7 to 7.7 million between 2010 to 2019, and from 2019 to 2021 decreased back to 6.2 million. Child and maternal malnutrition risks were generally below 600 thousand deaths except for low birth weight and short gestation childhood undernutrition and child growth failure that remained stable. There were no Level 3 metabolic risk factors.

### Variability of estimates

Over all estimates the median of R/M estimates from 2010-2023 was 0.8 (full range 0–3.8) for deaths and 0.7 for DALYs (full range 0.1–3.3). When limited to the latest four iterations (2017, 2019, 2021, 2023) the median R/M was 0.5 (full range 0–2.9) for deaths and 0.4 (full range 0–2.8) for DALYs. Among Level 1 risk categories, the median R/M was 1.0 for deaths and 0.9 for DALYs attributable to behavioral risks, 0.7 for deaths and 0.6 for DALYs attributable to environmental and occupational risks, and 0.5 and 0.6 for deaths and DALYs respectively attributable to metabolic risks. At Level 2, the highest R/M values were observed for other environmental risks (2.4 for deaths and 2.3 for DALYs), low physical activity (1.7 for deaths and 1.8 for DALYs), and intimate partner violence (1.1 for deaths and DALYs).

Across the 2010-2023 GBD iterations, the risk groups that had the highest proportion of factors with R/M>1, were behavioral risks (14/34, 41% for deaths and 17/34, 50% for DALYs) and environmental and occupational risks (6/24, 25% for deaths and 5/24. 21% for DALYs). The risk factors contributing to the high R/M for the behavioral risks’ death estimates were primarily dietary risks (7/16, 44%), and child and maternal malnutrition (3/8, 50%) (eTable 2). Risk factors with R/M above 1.5 for deaths and/or DALYs mainly consisted of behavioral risks such as dietary risks and child and maternal malnutrition (Table 1). When we limited the analysis to estimates from the four most recent iterations, the proportion of estimates with R/M>1 was 10/33 (30%) for behavioral risks’ death estimates (9/33, 27% for DALYs), 2/24 (8%) for environmental/occupational risks (2/24, 8% for DALYs) and 0/7 for metabolic risks (0/7 for DALYs).

### Comparisons for original and subsequently revised estimates for the same year

Across all 66 risk factors and 5100 total estimates from different years where revisions had been performed, median R/M was 0.5 for deaths and 0.4 for DALYs, but varied widely (0-2.9 for deaths and 0-2.9 for DALYs), while the interquartile range for deaths (0.2-1) and DALYs (0.2-0.8) was narrower.

145/675 (21%) risk factor estimates had R/M over 1 for deaths and 120/675 (18%) risk factor estimates had R/M over 1 for DALYs. The highest proportions of such large variability were seen for behavioral risk factors with R/M>1 in 108/356 (30%) of death estimates and 85/356 (24%) of DALY estimates. For environmental and metabolic risks, the same proportions were below or equal to 10% (eTable 3). The Level 2 risks that contributed most to the high proportion among the behavioral risks were dietary risks with a proportion of 72/176 (41%) for deaths and 54/176 (31%) for DALYs, child and maternal malnutrition with a proportion of 19/81 (23%) and 14/81 (17%) for deaths and DALYs, and low physical activity with a proportion of 5/11 (45%) and 5/11 (45%) for deaths and DALYs, respectively. The risk factors with R/M above 1.5 were diet high in sugar-sweetened beverages, in red meat and diet low in fruits, vegetables, nuts and seeds, seafood omega-3 fatty acids as well as low physical activity, Iron, Zinc and Vitamin A deficiency, chewing tobacco, lead exposure and other environmental risks (Figure 2A).

**Figure 2.**
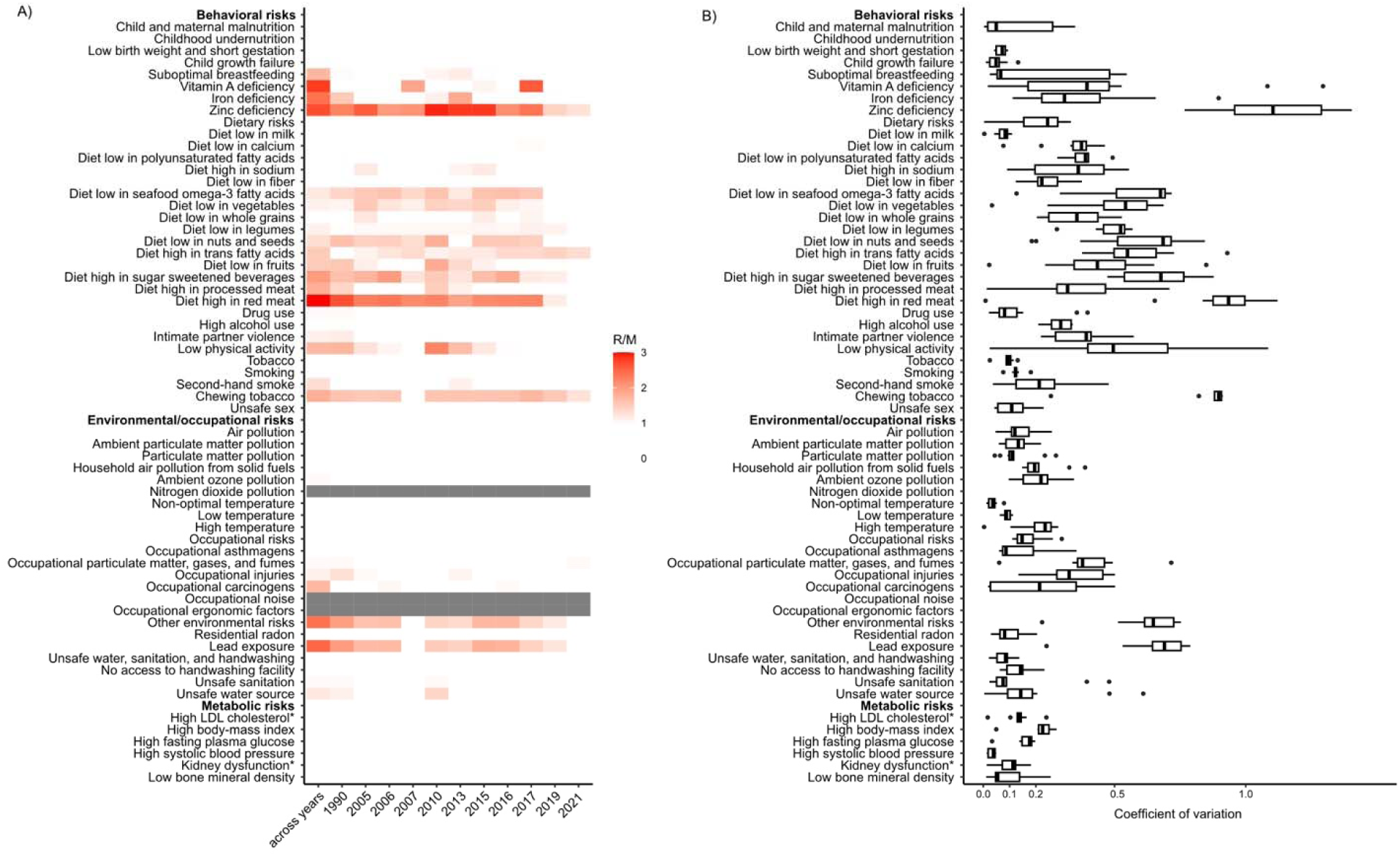
Variability in death estimates using the range-to-mean ratio (heatmap, A) and coefficient of variation (boxplot, B). In the heatmap (A) the first column shows variability across index years (e.g., GBD 2010 for 2010 through GBD 2023 for 2023), and subsequent columns show variability in matched-year estimates comparing original and revised values for the same calendar year. Note: nitrogen dioxide pollution, occupational noise, and ergonomic factors had no corresponding death estimates.

To validate our findings with the R/M approach we calculated the coefficient of variation showing that 336/675 (50%) of death estimates had a CV>0.2, and 127/675 (19%) had CV>0.5. They were mainly elevated among the behavioral and environmental/occupational risks (Figure 2B). Results for DALYs were similar (eFigure 3).

Figure 3 provides a plot of the Level 1 risk estimates across GBD iterations and years assessed as an illustration of the deviations for the same risk and year. The deviations were most prominent among environmental/occupational risks and behavioral risks. The same plots for deaths and DALYs of Level 2 risks are found in eFigure 4 and 5 respectively, in brief the same discrepancies across dietary risks, child and maternal malnutrition were observed as well as for air pollution for both deaths and DALYs.

**Figure 3.**
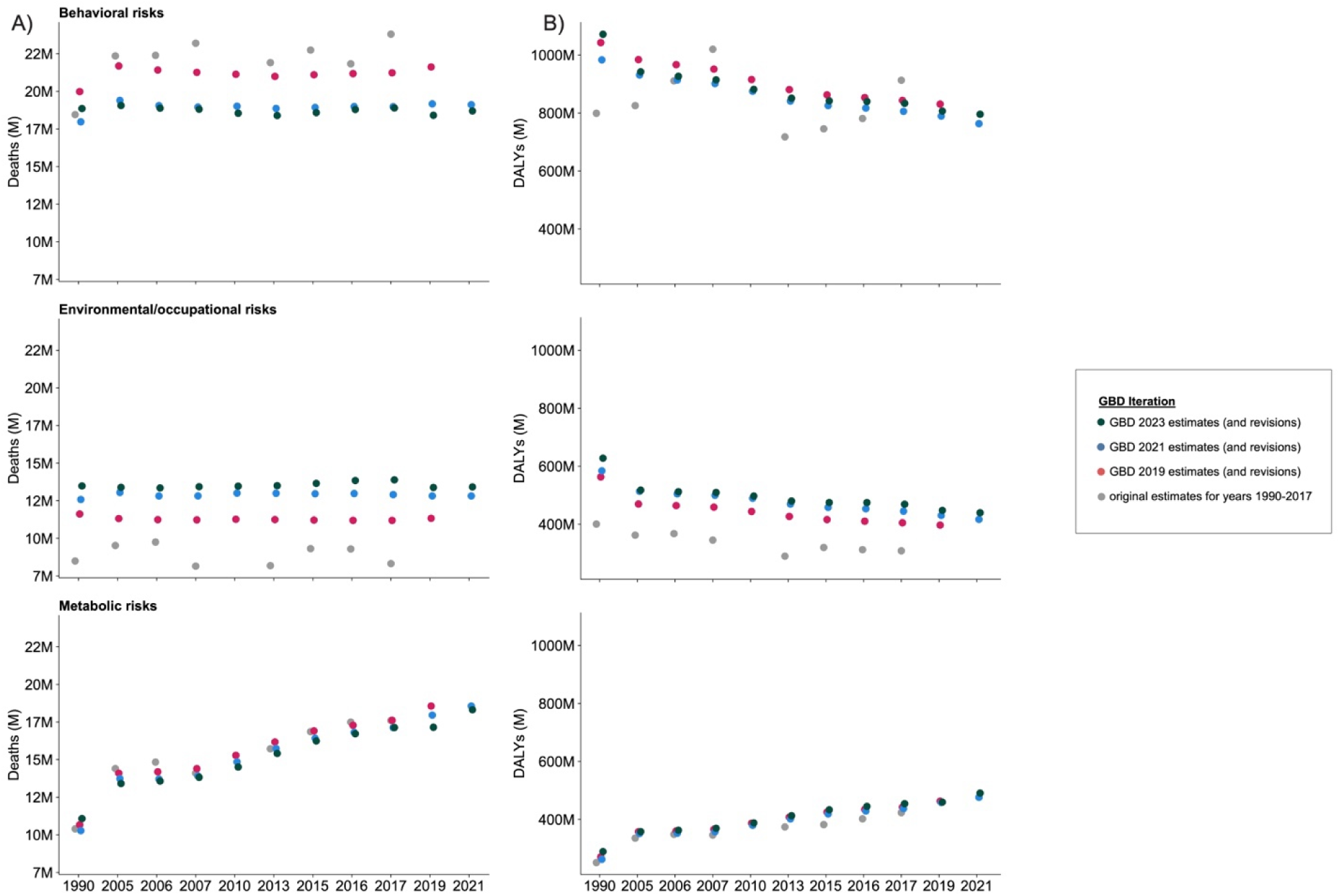
Matched risk year estimates for Level 1 risks for A) deaths and B) DALYs, revised in GBD 2023, 2021 and GBD 2019. *Note: GBD 2010 did not provide any Level 1 risk estimates*.

### Dietary risk factors and low physical activity: trends and fluctuations in estimates

The trends and fluctuations of the dietary risk factors and low physical activity are presented in detail as an illustrative example for highly debated factors. Fluctuations above 1 million deaths between iterations were common for diet high in sodium, low in nuts and seeds, low in whole grains and low physical activity for example. This pattern was also mirrored within the DALY estimates of the dietary risks (eFigure 6). Rankings of dietary risk factors also fluctuated substantially across iterations. For example, diet high in processed meat dropped from 8th to 13th in attributable deaths between GBD 2017 and GBD 2019, while diet high in red meat rose from 16th to 5th for both deaths and DALYs. No dietary risk factor maintained the same rank across more than three consecutive GBD iterations except for diet low in calcium for DALYs (eFigure 7).

### Dietary risk factors and low physical activity: consistency of estimates against previous 95%UIs

A comparison of all estimates for dietary risk factors and low physical activity, using gender, region-, and overarching cause-specific estimates, revealed that when comparing the GBD 2021 estimates to the 95%UIs of GBD 2019, approximately half of the revised dietary and low physical activity risk factor estimates fell outside the prior 95%UIs more than 25% of the time. The highest proportion of GBD 2021 death point estimates outside the GBD 2019 95%UI was observed for diet low in vegetables (882/1260, 70%), diet low in fruits (900/1260, 72%), diet high in red meat (1054/1260, 84%), diet low in seafood omega-3 fatty acids (1202/1260, 95%) and diet high in sugar-sweetened beverages (1209/1260, 96%). Similar proportions were observed for DALYs (Figure 4). The comparison of the GBD 2023 point estimate to GBD 2021 95%UI showed discrepancies in a majority of diet high in trans fatty acids estimates (992/1344, 77%) (Figure 4).

**Figure 4.**
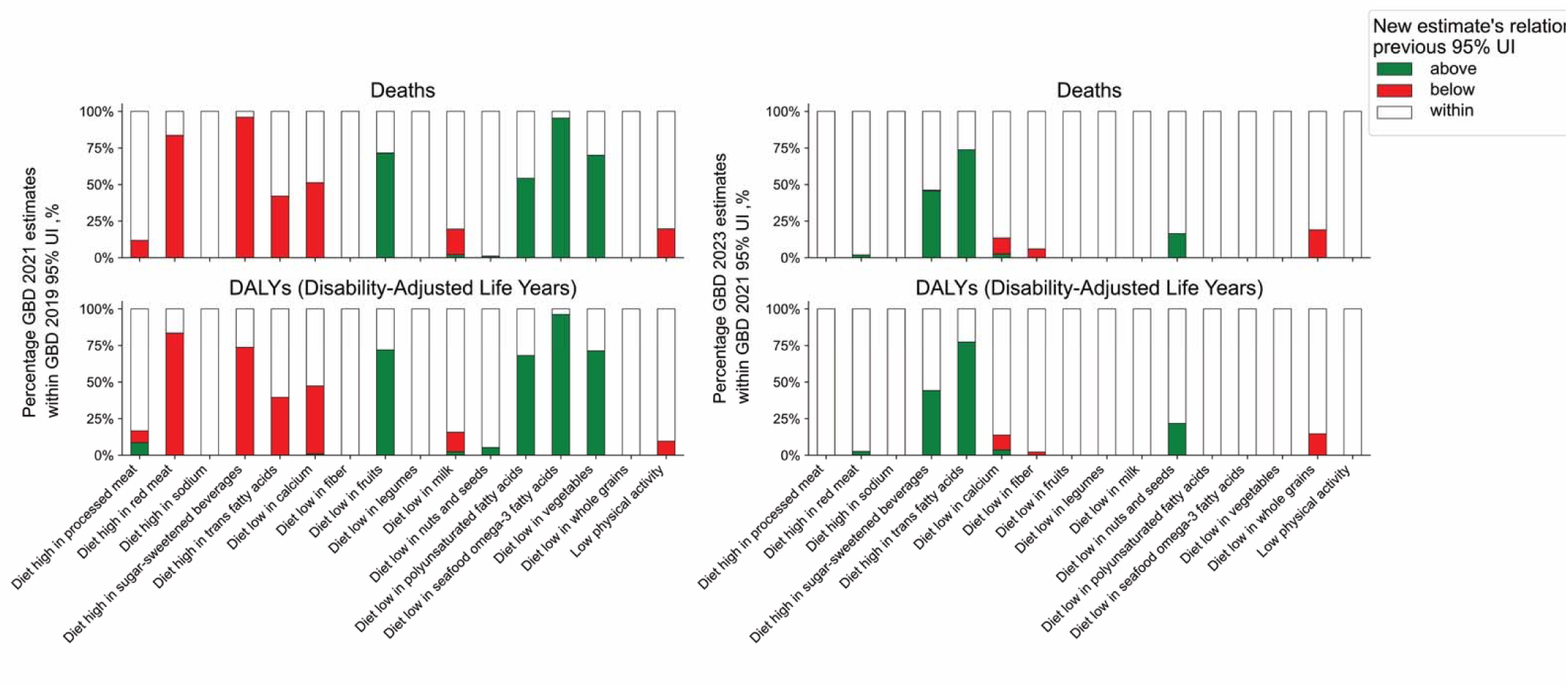
Consistency of dietary and low physical activity risk estimates between GBD 2019 and GBD 2021 and between GBD 2021 and GBD 2023. Left panels: Proportion of GBD 2021 point estimates above (red) below (green) and within (white) the corresponding GBD 2019 95% uncertainty interval (UI). Right panels: same comparison for GBD 2023 point estimates and GBD 2021 95%UIs. Risk factors are shown on the x-axis.

## DISCUSSION

In this analysis of the Global Burden of Disease (GBD) risk factor estimates through eight iterations spanning 2010 to 2023, we found substantial variability in mortality and DALY estimates. This was most prominent for behavioral risks such as diet and low physical activity. Even when restricting comparisons to the same calendar years, major differences between original and revised estimates persisted, suggesting that much of the variability reflects methodological changes rather than true changes in population health. In contrast, metabolic risk estimates were more stable across iterations.

Prior critiques^10–12,21^ have emphasized uncertainties in defining dietary exposures, thresholds for low physical activity, and reliance on heterogeneous observational data^22,23^. Our results provide empirical evidence that these uncertainties may manifest as unstable estimates across successive GBD publications. There are longstanding concerns about the reliability of risk factor epidemiology with vulnerability to confounding, measurement and sampling errors, as well as the effects of differential model specifications^24–27^.

The complexity inherent to computing population attributable fractions^28–31^ and the impact of different model specifications can yield divergent estimates when GBD is compared against other similar efforts at estimation of disease burden. Previously noted and debated examples include the more than twofold discrepancy between World Health Organization and GBD death estimates for malaria^32^ and the differences between the Non-Communicable Diseases Risk factor Collaboration (NCD-RisC) and the GBD estimates of obesity and overweight^33^. In situations such as traffic accidents, a comparison to national statistics in countries with high quality data has showed the GBD overestimated traffic accidents by 45% overall compared to the International Road Traffic Accident Database^34^.

Our study has several limitations. First, analyses were limited to published GBD estimates rather than primary data or replication of underlying models. Second, distinguishing true shifts in risk exposure from methodological artifacts is difficult; however, the degree of divergence between original and revised estimates for the same year suggests that modeling choices may play a major role. Third, early GBD iterations provided few overlapping years, limiting temporal comparisons. Risk factor definitions also evolved over time, although we have tried to minimize the impact of definition changes through harmonization (see eMethods).

Acknowledging these caveats, these findings suggest that GBD risk factor estimates that are increasingly used in policy decisions^35^ for behavioral risks, should be interpreted with caution and used with great care. Some of these estimates, in particular those referring to dietary factors, may be far more uncertain that their seeming precision suggests. Improved public health data may aid in improving the stability of global health risk factor estimates^31^.

## Data Availability

All code and data are publicly available on GitHub.

https://github.com/zavalis/pubs_meta/tree/main/gbd%20variability.

