## Supplementary Material for "Instability of Global Burden of Disease Estimates of Deaths and DALYs from Major Risk Factors"

**eMethods** **2**

**eFigure 1. Explanation of comparison of overlapping estimates for GBD 2019 and 2021.** **4**

**eFigure 3. Matched risk-year death estimates for level 2 risk factors.** **7**

**eFigure 4. Matched risk-year DALY estimates for level 2 risk factors.** **8**

**eFigure 5. Death (left) and DALY (right) estimates for the dietary risks across GBD iterations.** **9**

**eFigure 6. Changes in rank of dietary risks over time** **10**

**eTable 1. Years Analyzed in Global Burden of Disease (GBD) Iterations** **11**

**eTable 2. Levels of included Risk Factors from the Global Burden of Disease (GBD) Study** **12**

**eTable 3. Proportion of risks within level 1 and level 2 categories with range/median ratio above 1.** **13**

**eTable 4. Proportion of risks within level 1 and level 2 categories with range/median ratio above 1 for matched risk-year estimates.** **14**

**eMethods**

**Harmonization of risk factors across GBD iterations**

We harmonized the risk factors' labels to standardize terminology across GBD iterations. All changes involved risk factors that were equivalently defined but inconsistently named between versions. We chose to harmonize high total cholesterol and high LDL cholesterol despite the exposure differing in definition to achieve approximate equivalence of the risks.

**Spelling Differences**

- **Diet low in fibre** → *Diet low in fiber*
- **Occupational exposure to sulfuric acid** → *Occupational exposure to sulphuric acid*
- **Behavioral risks** → *Behavioural risks*
- **Second hand smoke** → *Second-hand smoke*
- **Secondhand smoke** → *Second-hand smoke*

**Specificity in wording (defined the same way)**

- **No handwashing with soap** → *No access to handwashing facility*
- **Physical inactivity and low physical activity** → *Low physical activity*
- **Short gestation for birth weight** → *Short gestation*
- **High blood pressure** → *High systolic blood pressure*
- **Alcohol use** → *High alcohol use*
- **Impaired kidney function** → *Kidney dysfunction*
- **Physiological risk factors** → *Metabolic risks*
- **Unsafe water, sanitation and handwashing** → *Unsafe water, sanitation, and handwashing*
- **Unimproved** → *Unsafe* (in water and sanitation contexts)
- **Tobacco smoking** → *Smoking*
- **Occupational risk factors for injuries** → *Occupational injuries*
- **Occupational risk factors** → *Occupational risks*
- **Childhood wasting** → *Child wasting*
- **Childhood stunting** → *Child stunting*
- **Diet low in omega 6 polyunsaturated fatty acids** → *Diet low in polyunsaturated fatty acids*
- **Diet suboptimal in calcium** → *Diet low in calcium*
- **Environmental risks** → *Environmental/occupational risks*
- **Environmental or occupational risks** → *Environmental/occupational risks*

**Differences in terms included but same semantic meaning**

(Synonymous terms and rewording for terminological alignment)

- **Tobacco smoking (including second-hand smoke)** → *Tobacco*
- **Tobacco smoke** → *Tobacco*
- **Low glomerular filtration rate**→Kidney dysfunction (although proteinuria was added in the definition of kidney dysfunction they correspond to the same level of risk and are non-overlapping as in cholesterol below, low GFR existed between GBD 2013 through GBD 2015 and Kidney dysfunction/Impaired kidney function GBD 2016 through GBD 2021)
- **High total cholesterol** → High LDL cholesterol (although they are not precisely the same the correspond to the same level of risks high total cholesterol existed between in GBD 2010 through GBD 2016 and high LDL cholesterol GBD 2017 through GBD 2021)

**Harmonization of levels across GBD iterations**

**Level 4 risks were removed**

- Non-exclusive breastfeeding
- Discontinued breastfeeding
- Childhood underweight
- Occupational exposure to asbestos
- Occupational exposure to arsenic
- Occupational exposure to benzene
- Occupational exposure to beryllium

- Occupational exposure to cadmium
- Occupational exposure to chromium
- Occupational exposure to diesel engine exhaust
- Occupational exposure to second-hand smoke
- Occupational exposure to formaldehyde
- Occupational exposure to nickel
- Occupational exposure to polycyclic aromatic hydrocarbons
- Occupational exposure to silica
- Occupational exposure to sulphuric acid
- Occupational exposure to trichloroethylene
- Child wasting
- Child stunting
- Child underweight
- Short gestation
- Low birth weight for gestation
- Low birth weight

**Abuse related risk factors with inconsistencies**

- Sexual abuse and violence
- Bullying victimization
- Sexual violence against children
- Sexual violence against children and bullying

**Alcohol and drug use (Level 2 risk)** was removed, and its constituents (namely high alcohol use and drug use) were treated as Level 2 risks instead as in GBD 2019 and GBD 2021.

#### eFigure 1. Explanation of comparison of overlapping estimates for GBD 2019 and 2021.

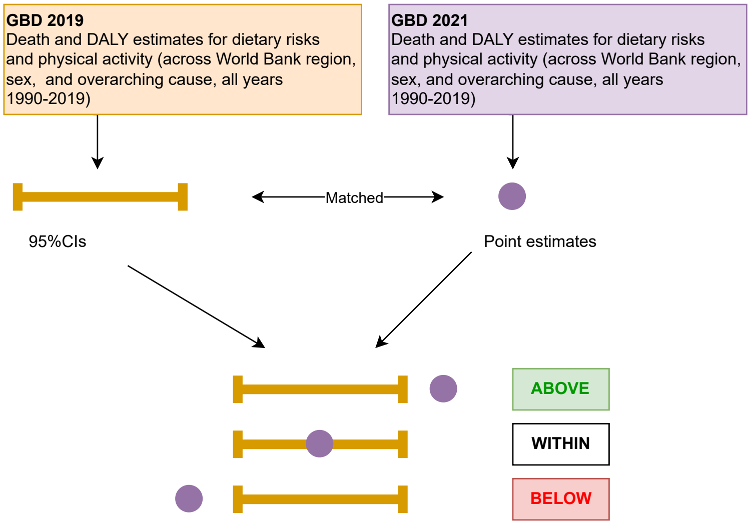


ABOVE means that the GBD 2021 estimate is above the GBD 2019 estimate's 95% confidence interval (CI). WITHIN means that the GBD 2021 estimate is within the GBD 2019 estimate's confidence interval. BELOW means that the GBD 2021 estimate is below the GBD 2019 estimate's confidence interval.

### eFigure 2. Estimated total deaths and DALYs attributable to Level 3 risk factors across GBD iterations.

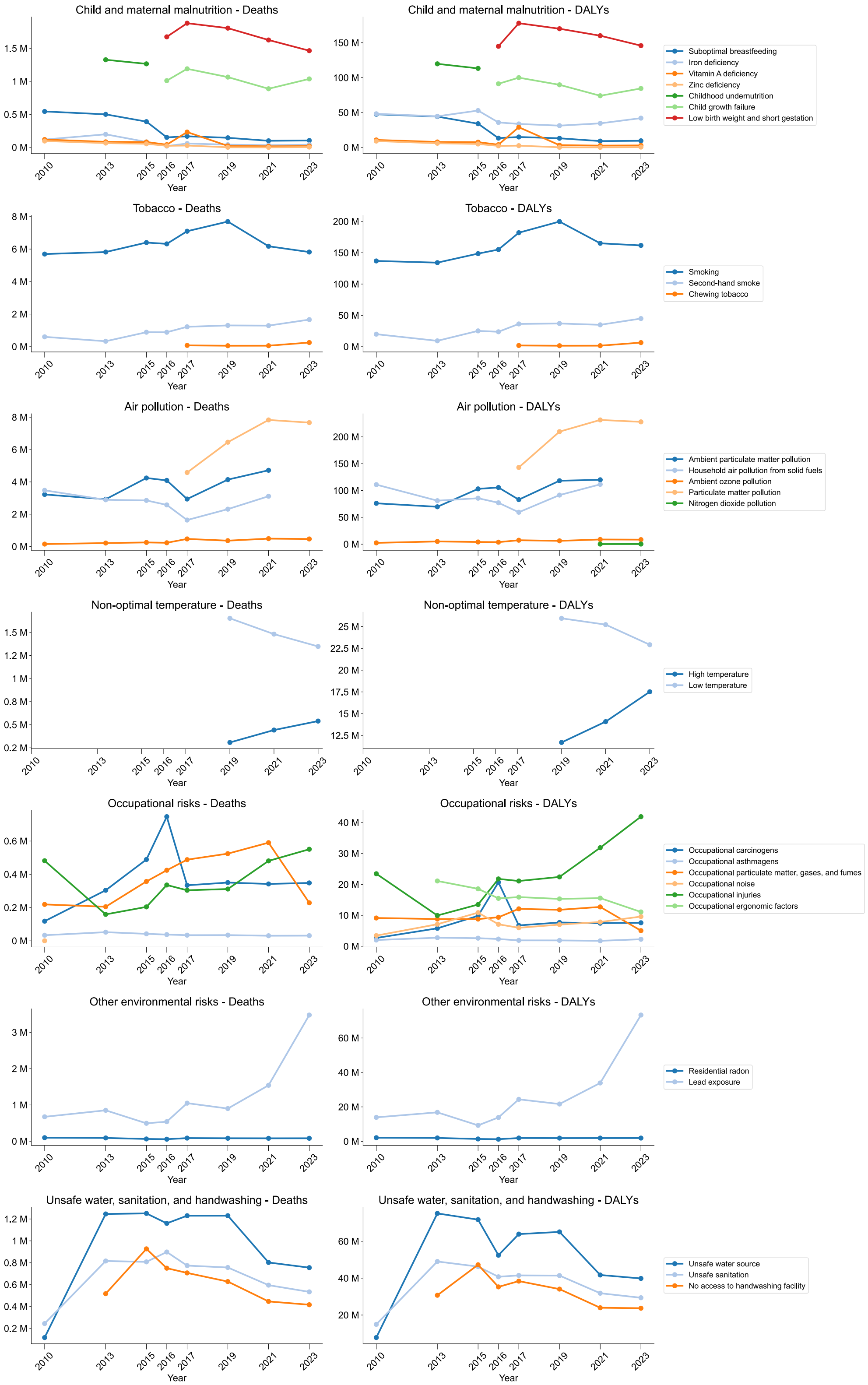

**eFigure 3. Variability in death using the range-to-mean ratio (A, heatmap) and coefficient of variation (B, boxplot).**

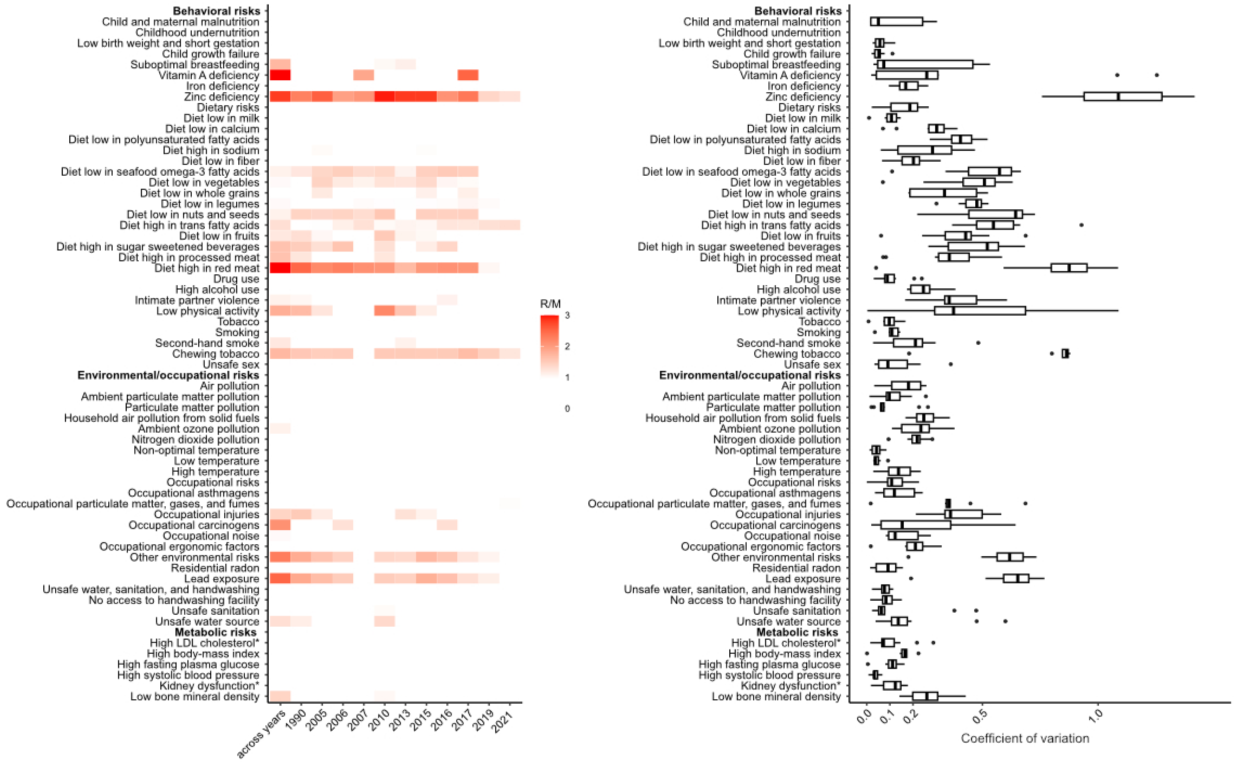


In A) the first column shows variability across index years (e.g., GBD 2010 for 2010 through GBD 2023 for 2023), and subsequent columns show variability in matched-year estimates comparing original and revised values for the same calendar year.

**eFigure 4. Matched risk-year death estimates for level 2 risk factors.**

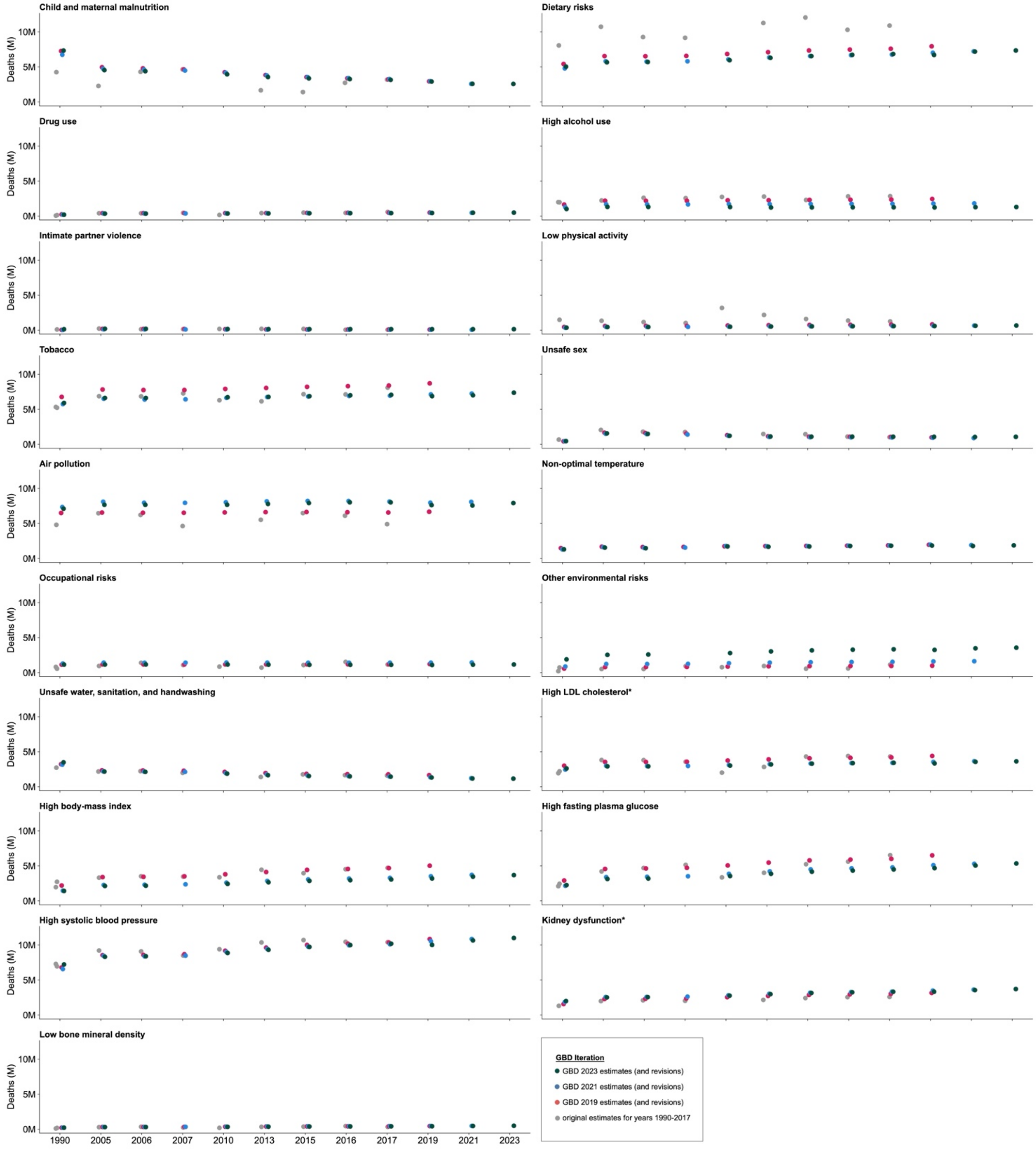

#### eFigure 5. Matched risk-year DALY estimates for level 2 risk factors.

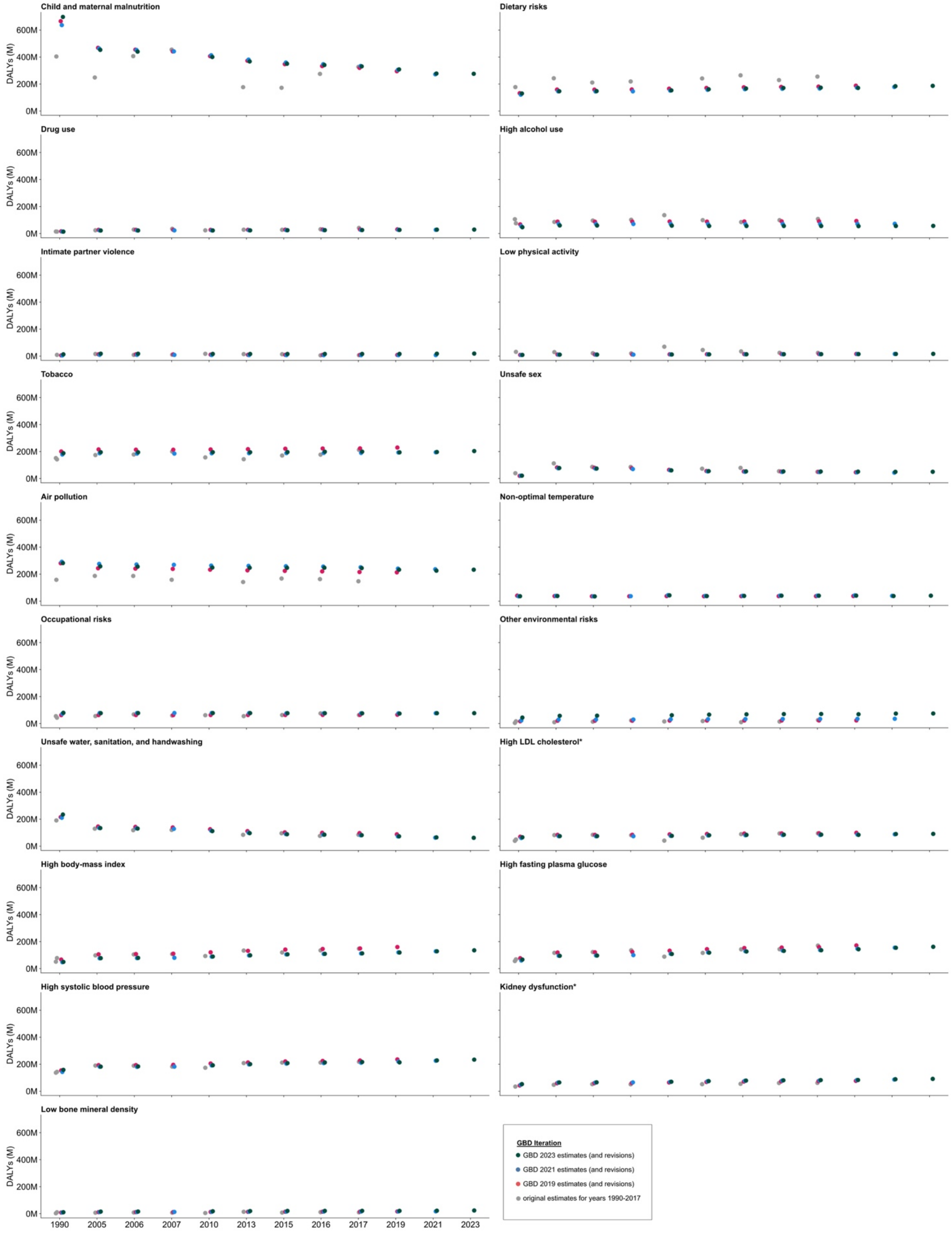

**eFigure 6. Death (left) and DALY (right) estimates for the dietary risks across GBD iterations.**

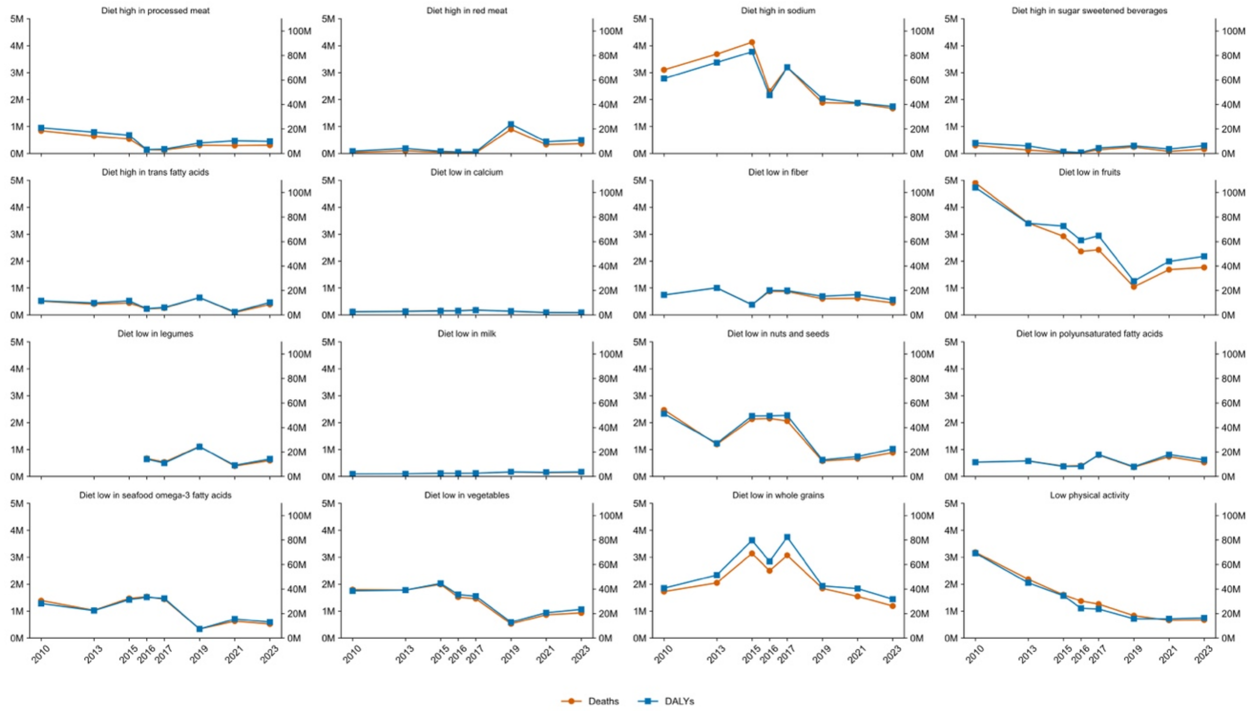


Y-axis is million (M) deaths on the left hand side and DALYs on the right, and the x-axis is GBD years.

**eFigure 7. Changes in rank of dietary risks over time**

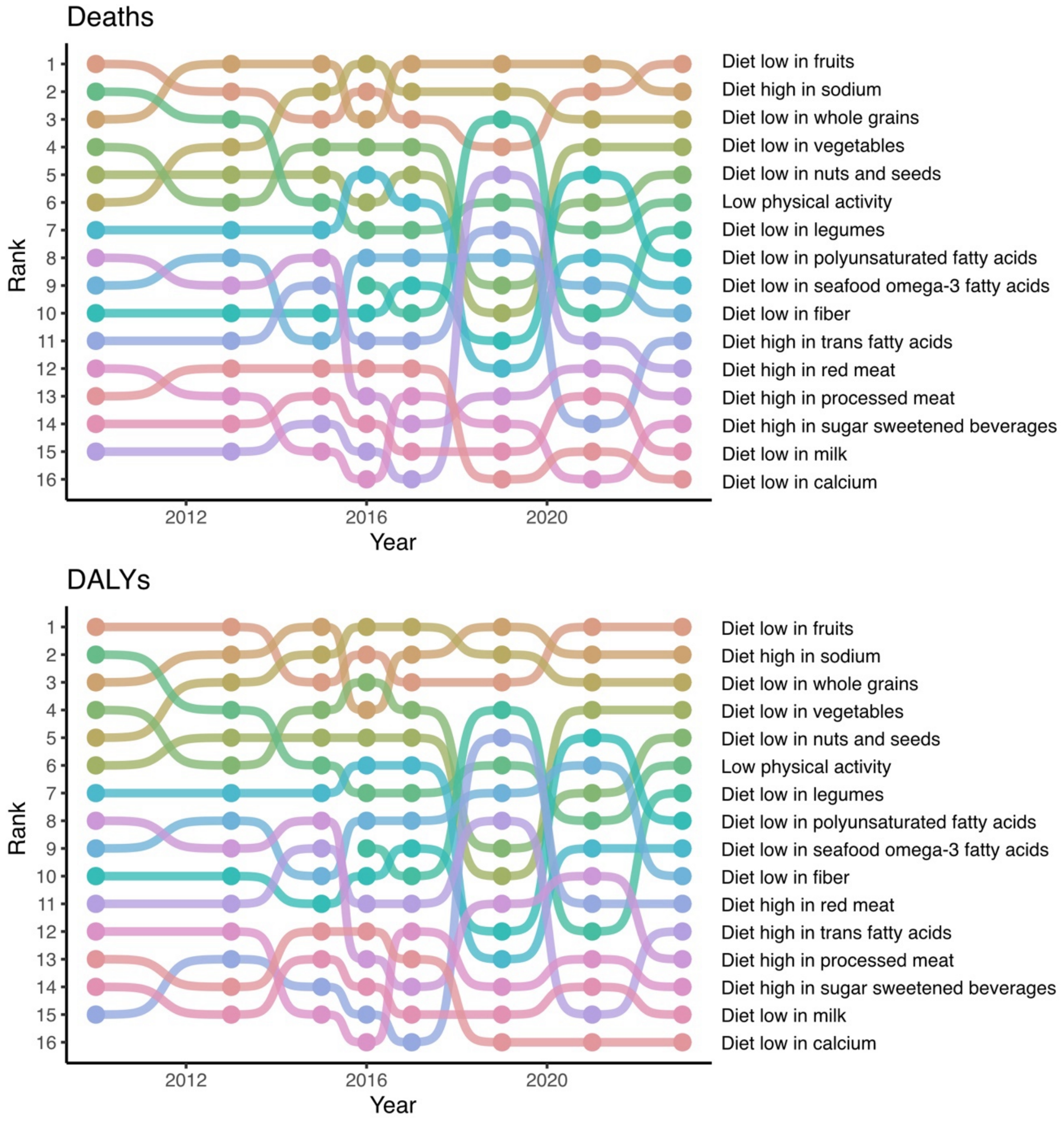

**eTable 1. Years Analyzed in Global Burden of Disease (GBD) Iterations**

*Level 1 risk factors were not estimated in GBD 2010.

| GBD iteration | Years estimated in the GBD iteration (utilized in this study) |
|---|---|
| 2010 | 1990, 2010* |
| 2013 | 1990, 2013 |
| 2015 | 2005, 2015 |
| 2016 | 2006, 2016 |
| 2017 | 2007, 2017 |
| 2019 | 1990, 2005, 2006, 2007, 2010, 2013, 2015, 2016, 2017, 2019<br>**Dietary risks:** 1980-2019 |
| 2021 | 1990, 2005, 2006, 2007, 2010, 2013, 2015, 2016, 2017, 2019, 2021<br>**Dietary risks:** 1980-2019 |
| 2023 | 1990, 2005, 2006, 2007, 2010, 2013, 2015, 2016, 2017, 2019, 2021<br>**Dietary risks:** 1990-2021 |

**eTable 2. Levels of included Risk Factors from the Global Burden of Disease (GBD) Study**

| | |
|---|---|
| **Behavioural risks** | |
| Child and maternal malnutrition | Suboptimal breastfeeding,<br>Iron deficiency,<br>Vitamin A deficiency,<br>Zinc deficiency,<br>Child and maternal malnutrition,<br>Childhood undernutrition, Child growth failure,<br>Low birth weight and short gestation |
| Dietary risks | Diet low in fruits, Diet low in vegetables, Diet low in legumes, Diet low in whole grains, Diet low in nuts and seeds, Diet low in fiber, Diet low in milk, Diet low in calcium, Diet low in seafood omega-3 fatty acids, Diet low in polyunsaturated fatty acids, Diet high in red meat, Diet high in processed meat, Diet high in sugar sweetened beverages, Diet high in trans fatty acids, Diet high in sodium |
| Drug use | |
| High alcohol use | |
| Intimate partner violence | |
| Low physical activity | |
| Tobacco | Smoking, Second-hand smoke, Chewing tobacco |
| Unsafe sex | |
| **Environmental/ occupational risks** | |
| Air pollution | Ambient particulate matter pollution, Household air pollution from solid fuels, Ambient ozone pollution, Particulate matter pollution, Nitrogen dioxide pollution |
| Non-optimal temperature | High temperature, Low temperature |
| Occupational risks | Occupational carcinogens, Occupational asthmagens, Occupational particulate matter, gases, and fumes, Occupational noise, Occupational injuries, Occupational ergonomic factors |
| Other environmental risks | Lead exposure, Residential radon |
| Unsafe water, sanitation, and handwashing | Unsafe water source, Unsafe sanitation, No access to handwashing facility |
| **Metabolic risks** | |
| High LDL cholesterol* | |
| High body-mass index | |
| High fasting plasma glucose | |
| High systolic blood pressure | |
| Kidney dysfunction* | |
| Low bone mineral density | |

**eTable 3. Proportion of risks within level 1 and level 2 categories with range/median ratio above 1.**

| Level 1 | Level 2 | Death estimates range/median> 1 (%) (n/N(%)) | DALY estimates range/median> 1 (n/N(%)) |
|---|---|---|---|
| **Behavioural risks** | **Overall behavioural risks estimate** | 17/34 (50) | 14/34 (41) |
| | Child and maternal malnutrition | 4/8 (50) | 3/8 (38) |
| | Dietary risks | 9/16 (56) | 7/16 (44) |
| | Drug use | 0/1 (0) | 0/1 (0) |
| | High alcohol use | 0/1 (0) | 0/1 (0) |
| | Intimate partner violence | 1/1 (100) | 1/1 (100) |
| | Low physical activity | 1/1 (100) | 1/1 (100) |
| | Behavioural risks | 0/1 (0) | 0/1 (0) |
| | Tobacco | 2/4 (50) | 2/4 (50) |
| | Unsafe sex | 0/1 (0) | 0/1 (0) |
| **Environmental and occupational risks** | **Overall risk environmental and occupational estimates** | 5/24 (21) | 6/24 (25) |
| | Air pollution | 0/6 (0) | 1/6 (17) |
| | Non-optimal temperature | 0/3 (0) | 0/3 (0) |
| | Occupational risks | 2/7 (29) | 2/7 (29) |
| | Environmental and occupational risks | 0/1 (0) | 0/1 (0) |
| | Other environmental risks | 2/3 (67) | 2/3 (67) |
| | Unsafe water, sanitation, and handwashing | 1/4 (25) | 1/4 (25) |
| **Metabolic risks** | **Overall metabolic risks estimate** | 0/7 (0) | 1/7 (014) |
| | High LDL cholesterol* | 0/1 (0) | 0/1 (0) |
| | High body-mass index | 0/1 (0) | 0/1 (0) |
| | High fasting plasma glucose | 0/1 (0) | 0/1 (0) |
| | High systolic blood pressure | 0/1 (0) | 0/1 (0) |
| | Kidney dysfunction* | 0/1 (0) | 0/1 (0) |
| | Low bone mineral density | 0/1 (0) | 1/1 (100) |
| | Metabolic risks | 0/1 (0) | 0/1 (0) |

**eTable 4. Proportion of risks within level 1 and level 2 categories with range/median ratio above 1 for matched risk-year estimates.**

| Level 1 | Level 2 | Death est. R/M>1 (%) (n/N(%)) | DALY est/ R/M>1 (n/N(%)) |
|---|---|---|---|
| **Behavioural risks** | **Overall behavioural risks estimate** | 111/356 (31) | 87/356 (24) |
| | Child and maternal malnutrition | 19/81 (23) | 14/81 (17) |
| | Dietary risks | 74/176 (42) | 55/176 (31) |
| | Drug use | 0/11 (0) | 0/11 (0) |
| | High alcohol use | 0/11 (0) | 0/11 (0) |
| | Intimate partner violence | 1/11 (9) | 1/11 (9) |
| | Low physical activity | 5/11 (45) | 5/11 (45) |
| | Tobacco | 12/44 (27) | 12/44 (27) |
| | Unsafe sex | 0/11 (0) | 0/11 (0) |
| **Environmental and occupational risks** | **Overall risk environmental and occupational estimates** | 24/253 (9) | 28/253 (11) |
| | Air pollution | 0/66 (0) | 0/66 (0) |
| | Non-optimal temperature | 0/33 (0) | 0/33 (0) |
| | Occupational risks | 2/77 (3) | 6/77 (8) |
| | Other environmental risks | 20/33 (61) | 20/33 (61) |
| | Unsafe water, sanitation, and handwashing | 2/44 (5) | 2/44 (5) |
| **Metabolic risks** | **Overall metabolic risks estimate** | 0/66 (0) | 0/66 (0) |
| | High LDL cholesterol* | 0/11 (0) | 0/11 (0) |
| | High body-mass index | 0/11 (0) | 0/11 (0) |
| | High fasting plasma glucose | 0/11 (0) | 0/11 (0) |
| | High systolic blood pressure | 0/11 (0) | 0/11 (0) |
| | Kidney dysfunction* | 0/11 (0) | 0/11 (0) |
| | Low bone mineral density | 0/11 (0) | 0/11 (0) |
